# HAI-VECT(SCD): AI-Humanoid Enabled Virtual Clinical Trial for Sickle Cell Disease

**DOI:** 10.1101/2023.10.17.23297152

**Authors:** WR Danter

## Abstract

**Background:** Sickle Cell Disease (SCD) remains a globally important disorder with limited therapeutic options. This study utilizes the advanced capabilities of the DeepNEU© platform v8.2 and aiHumanoid (Pat. Pend.) simulations to evaluate potential drug combinations for treating SCD, focusing on vaso-occlusive events (VOE) and associated secondary outcomes.

**Methods:** Using data from 25 virtual patients in each of six treatment groups, therapeutic responses to each treatment were investigated. The study evaluated the primary outcome of VOE and secondary outcomes, including HbA, HbF, Quality of Life (QoL), RBC hemolysis, and Pain. Treatment toxicities were also assessed across all dosage levels.

**Results:** The combination of Endari (L-glutamine powder) plus Crizanlizumab (a P-selectin antibody) demonstrated superior efficacy, with significant improvements in primary and secondary endpoints. This regimen, along with Voxelotor (a hemoglobin S polymerization inhibitor) plus Crizanlizumab, showed promising reductions in VOE, RBC hemolysis, and enhanced QoL scores. Notably, these results align with existing literature emphasizing the benefits of combination therapies in SCD management. Furthermore, aiHumanoid simulations indicated that these treatment combinations present lower cumulative multi-organoid toxicity, potentially translating to better patient outcomes and reduced healthcare costs.

**Conclusion:** AI-driven virtual clinical trials offer an innovative approach in evaluating drug combinations, presenting a robust case for the efficacy of Endari plus Crizanlizumab in managing SCD. The results warrant further research and real-world trials, potentially reshaping clinical guidelines for SCD treatment.

## Introduction

Sickle Cell Disease (SCD), affecting millions globally, is a debilitating genetic disorder resulting from a mutation in the β-globin gene, predominantly affecting populations of African, Mediterranean, Middle Eastern, and Indian descent [1]. It is often marked by severe complications including vaso-occlusive events (VOE), hemolytic anemia, and acute pain crises, leading to significant morbidity and mortality.

The current management of SCD crisis involves supportive measures such as Oxygen, IV fluids, analgesics, and blood transfusions for severe vaso-occlusive events like acute chest syndrome and stroke [2]. While hydroxyurea has been the standard for medical care due to its capacity to reduce VOE and the need for blood transfusions by inducing fetal hemoglobin (HbF) production, its use can be also accompanied by side effects such as myelosuppression, demanding the exploration of alternative therapies [3].

Several FDA-approved therapies such as Crizanlizumab (a P-selectin antibody)and Voxelotor (a hemoglobin S polymerization inhibitor) target different aspects of SCD pathophysiology. Crizanlizumab inhibits the interaction between sickle cells, leukocytes, and endothelial cells [4]. Voxelotor, investigated in the pivotal HOPE trial, showed efficacy in mitigating hemolysis and improving anemia [5]. A third drug, Endari (L-glutamine powder) manages SCD complications by modulating oxidative stress within sickle cells [6].

By harnessing the capabilities of aiHumanoid simulations to create virtual subjects of any target demographic, genotypic or phenotypic study population there is the real potential to expedite the clinical trial process, providing insights without the constraints of traditional trials. Given the complexities of SCD and the diversity in treatment modalities, this paper proposes a virtual Phase 1b/2 trial designed to evaluate all possible two-drug combinations of the approved drugs, with the primary endpoint focusing on the reduction of VOE. The goal of this trial is to optimize therapeutic strategies by combining drugs with different mechanisms of action, providing a comprehensive understanding of the potential synergies between these therapies in reducing VOE and improving the overall quality of life for SCD patients. Through the aiHumanoid simulations, this study takes a leap towards advancing personalized medicine for SCD patients. By simulating patient responses, we aim to identify the most promising drug combinations based on their efficacy and safety profiles, potentially revolutionizing our approach to SCD management.

The exploration and evaluation of these diverse therapeutic agents individually and in combination are imperative to enhance our understanding of their synergistic, additive, or potentially antagonistic interactions. This paper endeavors to critically assess these therapeutic agents, aiming to elucidate optimal therapeutic strategies and contribute to the evolving landscape of personalized medicine in SCD management.

## Methods

The current study (HAI-VECT(SCD)) extends our previous work using literature validated aiHumanoid simulations [7,8] as virtual patients. This virtual clinical trial was designed to compare the Efficacy, Toxicities and Quality of Life estimates for conventional drug therapy with hydroxyurea vs all unique two drug combinations of the four approved drugs for treating severe SCD: hydroxyurea, Voxelotor, L-glutamine powder and the monoclonal antibody Crizanlizumab.

### 1a. Updating the aiHumanoid Simulation to v8.2

The previous version 8.1 of the aiHumanoid [8] underwent modest updates to v8.2. The main differences are that the revised version includes new simulations of Quality of Life (Physical and Psychological) and important updates to Erythropoiesis, and RBC physiology subsystems. The total number of integrated organoid simulations remains at 21. Regarding literature validation of the SCD simulations, the same approach used in version 8.0 and 8.1 was applied to the new and updated simulations comprising v8.2. A detailed overview of this process is summarized in Appendix 1.

### 1b SCD Validation Profile in the aiHumanoid Simulations

To confirm the presence of SCD in the aiHumanoid simulations, the following 8 phenotypic features assembled from the literature were evaluated: HbA, HbF, Quality of Life-Physical, Quality of Life-Psychological, Reticulocytes, RBC Hemolysis, Pain, and Vaso-Occlusion Events (VOE) in addition to the HBB LOF mutation.

The final panel of SCD disease markers was determined by statistical analysis using a Z Score estimate of >3 when patient features profile changes from control were positive direction and <-5 when they were negative. The final panel of predictors for the healthy virtual patient profile and 25 virtual SCD patients were compared first with a correlation coefficient and a Bonferroni corrected p value <0.006 to determine whether SCD patient profiles were significantly different from the healthy controls.

As before [8] the group of 25 virtual SCD patient profiles was created by GPT4 from recent literature to cover a wide range of patient demographics and phenotypic characteristics than could be achieved with a single common patient simulation. All patient profiles include Age, Gender, Risk factors, Symptoms, HBB mutation status and VOE history. A representative group of virtual patient profiles is summarized in Appendix 2.

### 2. Rationale for the drug combinations

The validated aiHumanoid based SCD simulations were used to evaluate the therapeutic response of each of the 25 virtual patients to six unique two drug combinations. The current standard of care for serious SCD is generally supportive plus hydroxyurea. Hydroxyurea was approved by the FDA for the treatment of adults with Sickle Cell Anemia (SCA) on December 31, 1998, and it was later approved for use in children with SCD in 2017. The approval was based on evidence showing its efficacy in increasing fetal hemoglobin levels and reducing the frequency of vaso-occlusive crises.

This paper describes a virtual pivotal clinical trial demonstrating the efficacy of hydroxyurea in reducing the frequency of painful crises in patients with Sickle Cell Anemia, which contributed to its FDA approval.

The rationale for evaluating two-drug combinations of currently approved drugs for treating Sickle Cell Disease (SCD) is driven by the need for improved therapeutic strategies to alleviate the manifold complications of this chronic disorder [1]. SCD is characterized by several pathological processes including hemolysis, vaso-occlusion, inflammation, and endothelial dysfunction, and often requires a multifaceted treatment approach [9].

Each of the four approved drugs targets different aspects of the pathophysiology of SCD, with potential synergistic or additive effects when used in combination. For example, hydroxyurea induces fetal hemoglobin, which can inhibit the polymerization of sickle hemoglobin [3], while Crizanlizumab primarily acts to reduce the frequency of painful crises by blocking P-selectin-mediated multicellular adhesion [4]. Combining diverse mechanisms of action has the potential to treat the disease more comprehensively, potentially enhancing efficacy and improving patient outcomes in terms of reducing vaso-occlusive events, alleviating pain, and improving quality of life [5,6]. Evaluating these combinations in a rigorous scientific manner can provide valuable insights into the most effective multidrug regimens, paving the way for personalized treatment approaches and potentially unveiling novel therapeutic paradigms for managing SCD.]. The selection of the specific drug combinations was based on the individual drugs’ distinct modes of action, the potential to target different aspects of SCD pathophysiology simultaneously, and preliminary evidence from clinical settings suggesting enhanced therapeutic benefits when used in combination.

### 3. Study Design and Objectives

This is the first of its kind, aiHumanoid based multi-arm, dose-escalation virtual clinical trial. The primary objective is to determine the potential ability of unique two drug combinations to reduce vaso-occlusive events (VOE)compared with standard of care, hydroxyurea, in virtual patients with SCD. Important secondary outcomes include cumulative toxicity, pain relief, and impact on quality of life. An overview of the study design is presented in Table 1a.

**Table 1a:** SCD Virtual trial design overview.

| Treatment groups | 25%* | 50% | 75% | 100% | N** |
| --- | --- | --- | --- | --- | --- |
| Group 1: Endari (L-glutamine) plus Crizanlizumab | ✓ | ✓ | ✓ | ✓ | 25 |
| Group 2: hydroxyurea/HU plus Crizanlizumab | ✓ | ✓ | ✓ | ✓ | 25 |
| Group 3: hydroxyurea/HU plus Endari | ✓ | ✓ | ✓ | ✓ | 25 |
| Group 4: Voxelotor plus Crizanlizumab | ✓ | ✓ | ✓ | ✓ | 25 |
| Group 5: Voxelotor plus Endari | ✓ | ✓ | ✓ | ✓ | 25 |
| Group 6: Voxelotor plus hydroxyurea/HU | ✓ | ✓ | ✓ | ✓ | 25 |

\*Percent of maximum dose, \*\* Sample size in each group

The virtual patients: The profiles for 25 unique virtual patients diagnosed with SCD were synthesized by GPT-4, OpenAI’s advanced language model (September 2023 version) https://chat.openai.com/. GPT4 used its extensive training data encompassing medical literature, patient profiles, and related clinical information, to synthesize diverse representative patient backgrounds, histories, genetic markers, and disease stages. This trial design permits us to create a virtual clinical trial with 6 (groups)X4(doses)X25 virtual patients) the equivalent of 600 patients. The virtual patients used in this study serve as hypothetical but commonly encountered examples of SCD but do not represent real individuals or precise medical histories.

(Note: GPT-4, developed by OpenAI, is a state-of-the-art language model capable of generating human-like text based on extensive training data. In this study, it was employed to generate virtual patient profiles, harnessing its vast knowledge from medical literature to ensure the profiles’ accuracy and relevance.)

Inclusion Criteria:

1. Diagnosis of SCD: Confirmed diagnosis of homozygous sickle cell anemia (HbSS) or compound heterozygous sickle cell disease (HbSC, HbSβ^0, or HbSβ^+ thalassemia).
2. Age: Individuals aged 12 years or older.
3. History of VOE: Patients must have experienced at least one VOE in the 12 months prior to inclusion.
4. Data Availability: Availability of sufficient historical clinical data to enable meaningful analysis.
5. Stable Treatment Regimen: Participants should be on a stable treatment regimen for at least 3 months prior to inclusion, with no changes to any other concurrent medication for SCD intended to prevent VOEs.

Exclusion Criteria:

1. Other Hemoglobinopathies: Individuals with other hemoglobinopathies or medical conditions that might confound the study results.
2. Pregnancy or Lactation: Women who are pregnant, planning to become pregnant, or breastfeeding during the study period.
3. Recent Blood Transfusion: Patients who have received a blood transfusion within 30 days prior to the start of the study.
4. Renal or Hepatic Dysfunction: Individuals with significant renal or hepatic dysfunction that might impact drug metabolism or excretion.
5. Severe Concurrent Illness: Any severe concurrent medical condition or illness that, in the investigator’s judgment, could compromise participant safety or compliance, interfere with consent, study conduct, or interpretation of study results.
6. Prior use of Study Drugs: Patients with prior exposure to the study drugs in combination within 3 months before the start of the study.
7. Inability to Comply: Individuals deemed unable to comply with study procedures and requirements.

#### Treatment and Dose Escalation

##### Dosing

Given the unique nature of virtual SCD patients, it is possible to use all 25 patients in each of the six groups of the study. In each group, the virtual patients will receive increasing doses of a specified unique two drug combination. The specific doses for all groups will be first 25%, then 50%, next 75%, and finally 100% of maximal dose. Dose escalation from 25% through 100% percent of maximal dose will be assessed after the safety and preliminary efficacy of the previous dose level have been determined.

##### Safety Assessments

The safety of standard of care and novel combination treatments is assessed using organoid based cellular toxicity markers derived from the current literature. A previously defined optimized subset of 11 organoid toxicity markers [8] was used to evaluate toxicity of both treatment arms in all virtual patients. These organoid specific markers can be found in Appendix 3.

For the purposes of this study dose-limiting organoid specific toxicities (DLTs) are defined compared to conventional therapy with hydroxyurea. Here we define a DLT as any individual patient data point in any of the toxicity markers that is more than 2 standard deviations above the mean for the 25 virtual patients compared to hydroxyurea alone.

### 4. Assessment of Response to Therapy Using a Specific Biomarker Subset

The optimal composite biomarker profile of the 8 features defined above plus multiorgan toxicity was used to assess the response to therapy in simulated SCD patients. Changes in the levels of these biomarkers post-treatment with the two drug combinations, relative to the hydroxyurea treatment group, serve as indicators of therapeutic efficacy.

#### Simulation Analysis and Interpretation

The statistical tests chosen were based on the nature of the data and the objectives of this analysis. A paired two-tailed T-test with Bonferroni correction was employed to account for repeated measurements on the same virtual patients, while Cohen’s d was chosen to measure the true effect size, a critical component when assessing the clinical significance beyond mere statistical relevance.

Step 1: For evaluating efficacy, toxicity and, quality of life differences between combination treatment groups and the hydroxyurea group, a paired two tailed T test was employed and a Bonferroni corrected p value of 0.006 or less is required for statistical significance.

Step 2: To further evaluate differences between treatment arms, Cohen’s d [10,11] was used to estimate real treatment effects as small, medium, or large.

Step 3: In the event of disagreement between p values and Cohen’s d values, decisions were based on Cohen’s d value.

Step 4: Within group differences were conducted in an analogous manner except that:

Step 5: To assess individual patient treatment associated cumulative toxicity, a Z-test was used to evaluate where each of the 25 virtual patients from all treatment groups stood in the distribution of toxicities seen in the hydroxyurea group. Cumulative toxicity estimates >= 2 SD above the mean of the hydroxyurea group estimates was defined as a dose limiting toxicities (DLT).

#### Statistical Analysis

Null hypothesis: The novel two drug combinations do not result in any differences in treatment outcomes for virtual SCD patients when compared to the standard of care (hydroxyurea). Given that multiple comparisons being conducted, a Bonferroni correction was applied, making the adjusted p value for rejecting the null hypothesis p < 0.006.

Alternate hypothesis: At least one of the two drug combinations produces better treatment outcomes for virtual SCD patients when compared to standard of care with hydroxyurea.

To assess the response to therapy, levels of an optimized subset of biomarkers (see Results section) were measured both before and after treatment interventions in both arms. The two-tailed paired t-test was conducted for each biomarker to determine if there was a statistically significant difference (p<0.006) in their levels between pre-treatment post treatment results. In addition, Cohen’s d was calculated to estimate the true treatment effects.

## Results

In the present study, v8.2 (2023) of the DeepNEU database was used, which is characterized by upgrades to its most recent previous version v8.1, [8]. Firstly, v8.1 consisted of 7267 genotypic and phenotypic concepts that were linked through 67491 positive and negative concepts. On the other hand, v8.2 has 7347 concepts and 68148 nonzero causal relationships. This indicates that for every nonzero concept in the causal relationship matrix, there were ∼ 9.2 incoming and outgoing causal relationships.

Furthermore, v8.2 offered new validated simulations for Physical and Psychological quality of life (QofL). Erythropoiesis and red blood cell (RBC) physiology subsystems were also updated in the current version. The total number of integrated organoid simulations remains at 21. The previously implemented early stopping algorithm (v8.1) was retained in this updated version. For the purposes of this project, we chose to use the results after 16 iterations, as determined through a 3 valued moving average, which avoided any evidence of overfitting.

### Virtual patient demographics

The gender specific characteristics of the 25 virtual SCD patients are summarized in Table 1b. The data indicate that the patients are well matched. There was only one statistically significant difference between male and female attributes at the p = 0.05 level. The virtual male patients appear to be heavier than female subjects.

**Table 1b:** Virtual patient demographics by gender.

| Virtual Patient Feature | Male | Female | Total | p Value |
| --- | --- | --- | --- | --- |
| Gender | 13 | 12 | 25 | 1 |
| AGE (yrs)* | 34.46±6.7 | 28.5±5.75 | 31.6±4.69 | 0.199 |
| HBB | 13 | 12 | 25 | 1 |
| Hypertension | 8 | 5 | 13 | 0.341 |
| Obesity | 9 | 3 | 12 | 0.041 |
| VOE | 13 | 12 | 25 | 1 |
| * ± 95% confidence interval |  |  |  |  |

### The SCD feature profile

Data for the 25 virtual patients were generated and analyzed for predictive value by comparing the profile with that from an unaffected aiHumanoid. A Z test was used to identify a subset of 8 features best predicted the presence of SCD. A two tailed paired T test was then used to confirm the predictive value of the final profile. The original p value of 0.05 was modified for multiple comparisons using the Bonferroni correction. This process created a corrected p value of 0.006. Using this method 8 of the original 14 features were selected for inclusion in the final disease profile. These eight factors are HbA, HbF, QoL-Physical, QoL-Psychological, Reticulocytes, Hemolysis, Pain, and VOE. A summary of this analysis appears in Table 2.

**Table 2:**
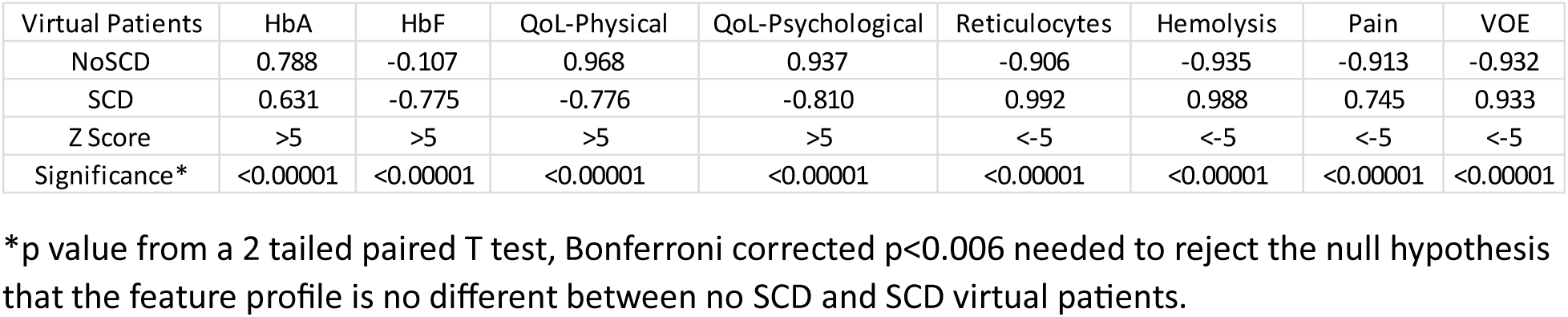
Optimized feature profile for confirming the presence of SCD.

| Virtual Patients | HbA | HbF | QoL-Physical | QoL-Psychological | Reticulocytes | Hemolysis | Pain | VOE |
| --- | --- | --- | --- | --- | --- | --- | --- | --- |
| NoSCD | 0.788 | -0.107 | 0.968 | 0.937 | -0.906 | -0.935 | -0.913 | -0.932 |
| SCD | 0.631 | -0.775 | -0.776 | -0.810 | 0.992 | 0.988 | 0.745 | 0.933 |
| Z Score | >5 | >5 | >5 | >5 | <-5 | <-5 | <-5 | <-5 |
| Significance* | <0.00001 | <0.00001 | <0.00001 | <0.00001 | <0.00001 | <0.00001 | <0.00001 | <0.00001 |

\*p value from a 2 tailed paired T test, Bonferroni corrected $p < 0.006$ needed to reject the null hypothesis that the feature profile is no different between no SCD and SCD virtual patients.

### Dose 1: 25% of Maximum Dose

The initial phase of formal testing started with a dose equivalent to 25% of the maximal dose for all drugs in both treatment arms. We utilized Cohen’s d to estimate the treatment effect. A two-tailed paired T-test was also used to determine the significance of the predicted differences observed between the hydroxyurea treatment group and the two drug combination groups. While T-test p-values provided insight, our primary metric for discerning a true treatment effect was Cohen’s d estimate, as recommended by sources including [10,11]. When Cohen’s d 95% Confidence Interval (CI) crosses 0 it is interpreted to mean that there is no effect.

For assessing toxicity, our analysis mirrored the above approach, but a Z-score was added to gauge hydroxyurea toxicities in comparison to the predicted toxicities for the other two drug treatment groups. A Z-score of ≥2 was set as a benchmark to identify potential toxic outliers in the two drug treatment groups. Using this approach, any treated patient with a Z-score of ≥2 was considered to have experienced a dose-limiting toxicity (DLT).

**Table 3a:** Efficacy vs hydroxyurea at 25% of maximum dose based on Cohen’s d +/- 95% CI.

| Treatment Option | HbA | HbF | QoFL-Physical | QoFL-Psychological | Reticulocytes | Hemolysis | Pain | VOE |
| --- | --- | --- | --- | --- | --- | --- | --- | --- |
| Endari plus Crizanlizumab | 1.62 | -0.22 | 0.19 | 0.19 | 0.00 | -0.01 | -0.08 | -0.02 |
| hydroxyurea/HU plus Crizanlizumab | -0.11 | 0.23 | 0.08 | 0.10 | -0.11 | -0.10 | -0.16 | -0.60 |
| hydroxyurea/HU plus Endari | -0.11 | 0.03 | 0.02 | 0.03 | -0.28 | -0.22 | -0.07 | -0.18 |
| Voxelotor plus Crizanlizumab | 1.64 | -0.25 | 0.26 | 0.31 | -0.28 | -0.24 | -0.19 | -0.49 |
| Voxelotor plus Endari | 1.49 | -0.32 | 0.16 | 0.21 | -0.43 | -0.24 | -0.01 | -0.04 |
| Voxelotor plus hydroxyurea/HU | -0.47 | 0.00 | 0.01 | 0.01 | -0.34 | -0.11 | -0.05 | -0.37 |

Summary: The combination of hydroxyurea plus Crizanlizumab at 25% of maximum dose produced a small to medium beneficial treatment effect on VOE based on Cohen’s d values (95% CI: -0.61+/- 0.34).

**Table 3b:** Cohen’s d +/- 95% CI for organoid specific toxicities compared to hydroxyurea at 25% of maximum dose.

| Treatment Option | Bone Marrow | Cardiac | CNS | CumTox | GI Tox | Immune | Liver Tox | Lung | Renal | Reproduction | Skin |
| --- | --- | --- | --- | --- | --- | --- | --- | --- | --- | --- | --- |
| Endari plus Crizanlizumab | -0.455 | -0.152 | -0.849 | -0.358 | 0.019 | 0.104 | -0.149 | -0.344 | 0.017 | -0.048 | 0.054 |
| Hydroxyurea/HU plus Crizanlizumab | 0.079 | -0.034 | -0.233 | -0.074 | -0.095 | -0.168 | -0.047 | -0.031 | -0.156 | -0.004 | -0.100 |
| Hydroxyurea/HU plus Endari | 0.074 | -0.002 | -0.075 | -0.060 | -0.009 | -0.039 | -0.304 | -0.131 | -0.091 | 0.004 | -0.219 |
| Voxelotor plus Crizanlizumab | -0.283 | -0.144 | -0.807 | -0.253 | 0.039 | 0.145 | 0.088 | -0.065 | 0.085 | -0.061 | 0.518 |
| Voxelotor plus Endari | -0.320 | -0.140 | -0.699 | -0.243 | 0.135 | 0.243 | -0.280 | -0.308 | 0.181 | -0.036 | 0.329 |
| Voxelotor plus Hydroxyurea/HU | 0.164 | -0.009 | -0.013 | -0.010 | -0.008 | -0.003 | -0.119 | -0.045 | -0.019 | -0.005 | -0.019 |

Summary: Based on Cohen’s d there is a trend toward a reduction in cumulative organoid toxicities involving multiple drug combinations. These Cohen’s d values suggest a trivial to small-medium beneficial effect across the drug combinations at 25% of maximum dose. The most effective combination at reducing cumulative toxicities appears to be Endari plus Crizanlizumab based on Cohen‘s d value (95% CI: -0.36 +/- 0.20).

**Table 3c:**
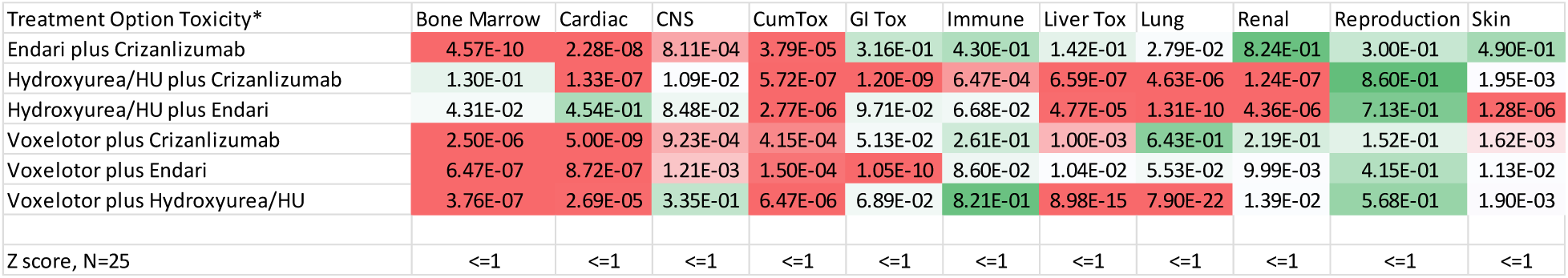
p values and Z scores for organoid specific toxicities compared to hydroxyurea at 25% of maximum dose.

| Treatment Option Toxicity* | Bone Marrow | Cardiac | CNS | CumTox | GI Tox | Immune | Liver Tox | Lung | Renal | Reproduction | Skin |
| --- | --- | --- | --- | --- | --- | --- | --- | --- | --- | --- | --- |
| Endari plus Crizanlizumab | 4.57E-10 | 2.28E-08 | 8.11E-04 | 3.79E-05 | 3.16E-01 | 4.30E-01 | 1.42E-01 | 2.79E-02 | 8.24E-01 | 3.00E-01 | 4.90E-01 |
| Hydroxyurea/HU plus Crizanlizumab | 1.30E-01 | 1.33E-07 | 1.09E-02 | 5.72E-07 | 1.20E-09 | 6.47E-04 | 6.59E-07 | 4.63E-06 | 1.24E-07 | 8.60E-01 | 1.95E-03 |
| Hydroxyurea/HU plus Endari | 4.31E-02 | 4.54E-01 | 8.48E-02 | 2.77E-06 | 9.71E-02 | 6.68E-02 | 4.77E-05 | 1.31E-10 | 4.36E-06 | 7.13E-01 | 1.28E-06 |
| Voxelotor plus Crizanlizumab | 2.50E-06 | 5.00E-09 | 9.23E-04 | 4.15E-04 | 5.13E-02 | 2.61E-01 | 1.00E-03 | 6.43E-01 | 2.19E-01 | 1.52E-01 | 1.62E-03 |
| Voxelotor plus Endari | 6.47E-07 | 8.72E-07 | 1.21E-03 | 1.50E-04 | 1.05E-10 | 8.60E-02 | 1.04E-02 | 5.53E-02 | 9.99E-03 | 4.15E-01 | 1.13E-02 |
| Voxelotor plus Hydroxyurea/HU | 3.76E-07 | 2.69E-05 | 3.35E-01 | 6.47E-06 | 6.89E-02 | 8.21E-01 | 8.98E-15 | 7.90E-22 | 1.39E-02 | 5.68E-01 | 1.90E-03 |
| Z score, N=25 | <=1 | <=1 | <=1 | <=1 | <=1 | <=1 | <=1 | <=1 | <=1 | <=1 | <=1 |

Summary: Importantly, no individual organoid toxicity measurement including the cumulative organoid toxicity estimate deviated by more than one standard deviation from the mean predicted toxicity for the hydroxyurea group.

Assessment: The analysis above reveals possible small reductions in anticipated organoid toxicities between the treatment groups and hydroxyurea. In the absence of evidence of DLT, we proceeded to increase the dosage for all drug combinations to 50% of the maximal dose in all groups.

### Dose 2: 50% of maximum dose

The analysis of all patient data for all treatment groups was conducted in an identical manner to that outlined above for Dose 1 Efficacy and Toxicity.

**Table 4a:** Efficacy vs hydroxyurea at 50% of maximum dose based on Cohen’s d.

| Treatment Option | HbA | HbF | QoL-Physical | QoL-Psychological | Reticulocytes | Hemolysis | Pain | VOE |
| --- | --- | --- | --- | --- | --- | --- | --- | --- |
| Endari plus Crizanlizumab | 1.156 | -0.175 | 0.501 | 0.511 | -0.108 | -0.672 | -0.612 | -0.525 |
| hydroxyurea/HU plus Crizanlizumab | 0.006 | 0.265 | 0.323 | 0.325 | -0.136 | -0.180 | -0.318 | -0.401 |
| hydroxyurea/HU plus Endari | -0.082 | -0.037 | 0.060 | 0.054 | -0.314 | -0.315 | -0.095 | -0.164 |
| Voxelotor plus Crizanlizumab | 1.007 | -0.281 | 0.435 | 0.445 | -0.196 | -0.464 | -0.475 | -0.509 |
| Voxelotor plus Endari | 0.853 | -0.512 | 0.250 | 0.245 | -0.353 | -0.333 | -0.217 | -0.102 |
| Voxelotor plus hydroxyurea/HU | -0.053 | 0.017 | 0.056 | 0.056 | -0.356 | -0.267 | -0.098 | -0.300 |

Summary: The combination of Endari plus Crizanlizumab at 50% of maximum dose produced a medium beneficial treatment effect on VOE with a Cohen’s d value of -0.53. A close second was the combination of Voxelotor plus Crizanlizumab with Cohen’s d value of -0.51.

**Table 4b:** Cohen’s d for organoid specific toxicities compared to hydroxyurea at 50% of maximum dose.

| Treatment Option Toxicity* | Bone Marrow | Cardiac | CNS | CumTox | GI Tox | Immune | Liver Tox | Lung | Renal | Reproduction | Skin |
| --- | --- | --- | --- | --- | --- | --- | --- | --- | --- | --- | --- |
| Endari plus Crizanlizumab | -0.938 | -0.246 | -1.203 | -0.719 | -0.210 | -0.624 | -0.318 | -0.335 | -0.398 | -0.155 | -0.225 |
| Hydroxyurea/HU plus Crizanlizumab | 0.209 | -0.094 | -0.587 | -0.225 | -0.100 | -0.491 | -0.132 | -0.017 | -0.237 | 0.160 | -0.148 |
| Hydroxyurea/HU plus Endari | 0.300 | 0.004 | -0.086 | -0.149 | -0.041 | -0.188 | -0.576 | -0.115 | -0.123 | -0.003 | -0.182 |
| Voxelotor plus Crizanlizumab | -0.622 | -0.215 | -1.115 | -0.472 | -0.021 | -0.362 | 0.091 | 0.070 | -0.146 | -0.161 | 0.051 |
| Voxelotor plus Endari | -0.502 | -0.190 | -0.762 | -0.391 | 0.191 | 0.151 | -0.370 | -0.283 | 0.162 | -0.141 | 0.017 |
| Voxelotor plus Hydroxyurea/HU | 0.484 | -0.036 | -0.067 | -0.046 | -0.013 | -0.086 | -0.165 | -0.170 | -0.052 | -0.004 | -0.066 |

Summary: Based on Cohen’s d there is a trend toward a reduction in cumulative organoid toxicities involving multiple drug combinations. These Cohen’s d values suggest a trivial to small-medium beneficial effect for most of the drug combinations at 50% of maximum dose. The most effective combination at reducing cumulative toxicities appears to be Endari plus Crizanlizumab with a Cohen ‘s d value of -0.72 (95% CI: -0.72+/-0.41). This suggests at least a medium beneficial effect on cumulative toxicity.

**Table 4c:** p values and Z scores for organoid specific toxicities compared to hydroxyurea at 50% of maximum dose.

| Treatment Option Toxicity* | Bone Marrow | Cardiac | CNS | CumTox | GI Tox | Immune | Liver Tox | Lung | Renal | Reproduction | Skin |
| --- | --- | --- | --- | --- | --- | --- | --- | --- | --- | --- | --- |
| Endari plus Crizanlizumab | 1.56E-14 | 2.78E-07 | 3.19E-05 | 1.09E-05 | 4.70E-02 | 2.50E-02 | 5.50E-03 | 6.24E-03 | 4.81E-02 | 2.00E-02 | 3.90E-03 |
| Hydroxyurea/HU plus Crizanlizumab | 9.09E-05 | 1.33E-07 | 1.53E-03 | 1.56E-05 | 1.04E-06 | 3.04E-02 | 2.06E-06 | 1.84E-11 | 3.83E-06 | 1.20E-05 | 3.18E-04 |
| Hydroxyurea/HU plus Endari | 4.55E-06 | 4.27E-01 | 1.24E-01 | 5.64E-09 | 3.04E-04 | 9.68E-03 | 3.59E-07 | 1.23E-10 | 3.70E-07 | 7.94E-01 | 3.41E-05 |
| Voxelotor plus Crizanlizumab | 4.14E-10 | 1.13E-10 | 2.84E-05 | 1.29E-05 | 7.15E-01 | 1.58E-01 | 4.92E-08 | 5.40E-01 | 3.04E-01 | 5.12E-03 | 1.87E-01 |
| Voxelotor plus Endari | 2.61E-08 | 5.16E-07 | 7.91E-04 | 1.97E-05 | 9.92E-10 | 3.28E-01 | 8.64E-04 | 3.61E-02 | 1.16E-02 | 1.56E-02 | 3.41E-01 |
| Voxelotor plus Hydroxyurea/HU | 5.50E-13 | 1.01E-07 | 8.68E-02 | 4.00E-04 | 1.77E-02 | 3.32E-01 | 4.01E-07 | 6.68E-28 | 8.25E-06 | 7.40E-01 | 3.31E-03 |
| Z score, N=25 | <=1 | <=1 | <=1 | <=1 | <=1 | <=1 | <=1 | <=1 | <=1 | <=1 | <=1 |
| * p value relative to Hydroxyurea Toxicity |  |  |  |  |  |  |  |  |  |  |  |

Summary: Again, no individual organoid toxicity measurement including the cumulative organoid toxicity estimate deviated by more than one standard deviation from the mean predicted toxicity for the hydroxyurea group.

Assessment: In the absence of evidence of DLT, and the indication of medium reduction in VOE, we proceeded to increase the dosage for all drug combinations to 75% of the maximal dose in all groups.

### Dose 3: 75% of maximum dose

The analysis of all patient data for all treatment groups was conducted in an identical manner to that outlined above for Dose 1 and Dose 2 Efficacy and Toxicity.

**Table 5a:** Efficacy vs hydroxyurea at 75% of maximum dose based on Cohen’s d.

| Treatment Option | HbA | HbF | QoL-Physical | QoL-Psychological | Reticulocytes | Hemolysis | Pain | VOE |
| --- | --- | --- | --- | --- | --- | --- | --- | --- |
| Endari plus Crizanlizumab | 0.600 | -0.019 | 0.249 | 0.245 | -0.064 | -1.059 | -0.370 | -0.691 |
| hydroxyurea/HU plus Crizanlizumab | 0.112 | 0.292 | 0.185 | 0.184 | -0.137 | -0.456 | -0.255 | -0.368 |
| hydroxyurea/HU plus Endari | -0.050 | 0.112 | 0.079 | 0.075 | -0.272 | -0.429 | -0.122 | -0.191 |
| Voxelotor plus Crizanlizumab | 0.354 | -0.160 | 0.180 | 0.175 | -0.171 | -0.641 | -0.244 | -0.401 |
| Voxelotor plus Endari | 0.404 | -0.389 | 0.086 | 0.080 | -0.252 | -0.416 | -0.057 | 0.036 |
| Voxelotor plus hydroxyurea/HU | -0.028 | 0.082 | 0.045 | 0.041 | -0.219 | -0.358 | -0.088 | -0.201 |

Summary: The combination of Endari plus Crizanlizumab at 75% of maximum dose produced a medium beneficial treatment effect on VOE with a Cohen’s d value of -0.69 (95% CI: -0.69+/-0.39).

**Table 5b:** Cohen’s d for organoid specific toxicities compared to hydroxyurea at 75% of maximum dose.

| Treatment Option Toxicity* | Bone Marrow | Cardiac | CNS | CumTox | GI Tox | Immune | Liver Tox | Lung | Renal | Reproduction | Skin |
| --- | --- | --- | --- | --- | --- | --- | --- | --- | --- | --- | --- |
| Endari plus Crizanlizumab | -0.814 | -0.472 | -0.729 | -0.819 | -0.571 | -0.486 | -0.423 | -0.226 | -0.818 | -0.512 | -0.368 |
| Hydroxyurea/HU plus Crizanlizumab | 0.043 | -0.195 | -0.270 | -0.289 | -0.208 | -0.243 | -0.175 | -0.015 | -0.491 | 0.170 | -0.160 |
| Hydroxyurea/HU plus Endari | 0.192 | 0.020 | -0.063 | -0.175 | -0.129 | -0.148 | -0.349 | -0.080 | -0.211 | 0.164 | -0.190 |
| Voxelotor plus Crizanlizumab | -0.578 | -0.429 | -0.497 | -0.423 | -0.144 | -0.236 | 0.124 | 0.182 | -0.310 | -0.577 | 0.142 |
| Voxelotor plus Endari | -0.543 | -0.325 | -0.301 | -0.310 | 0.267 | 0.042 | -0.118 | -0.153 | 0.127 | -0.498 | 0.150 |
| Voxelotor plus Hydroxyurea/HU | 0.190 | -0.033 | -0.053 | -0.059 | -0.006 | -0.008 | -0.082 | -0.091 | -0.079 | -0.016 | -0.061 |

Summary: Based on Cohen’s d there is a strong trend toward a reduction in cumulative organoid toxicities involving multiple drug combinations. These Cohen’s d values suggest a small to large beneficial effect for most of the drug combinations compared to hydroxyurea at 75% of maximum dose. The most effective combination at reducing cumulative toxicities appears to be Endari plus Crizanlizumab with a Cohen ‘s d value of -0.82 (95% CI: -0.82+/-0.47). This suggests a large effect on reducing cumulative toxicity compared to hydroxyurea.

**Table 5c:** p values and Z scores for organoid specific toxicities compared to hydroxyurea at 75% of maximum dose.

| Treatment Option Toxicity* | Bone Marrow | Cardiac | CNS | CumTox | GI Tox | Immune | Liver Tox | Lung | Renal | Reproduction | Skin |
| --- | --- | --- | --- | --- | --- | --- | --- | --- | --- | --- | --- |
| Endari plus Crizanlizumab | 2.19E-16 | 6.37E-08 | 6.24E-07 | 4.76E-06 | 3.11E-03 | 2.01E-03 | 1.79E-02 | 5.45E-03 | 1.49E-03 | 1.67E-04 | 2.17E-02 |
| Hydroxyurea/HU plus Crizanlizumab | 7.87E-02 | 1.87E-12 | 9.07E-05 | 6.33E-07 | 2.11E-03 | 1.13E-02 | 1.98E-05 | 3.91E-02 | 1.33E-06 | 6.84E-03 | 2.61E-05 |
| Hydroxyurea/HU plus Endari | 3.95E-07 | 1.82E-02 | 6.81E-03 | 1.39E-13 | 8.18E-05 | 2.13E-03 | 8.91E-15 | 1.47E-11 | 1.10E-08 | 4.67E-03 | 8.07E-08 |
| Voxelotor plus Crizanlizumab | 1.37E-11 | 2.00E-11 | 7.91E-06 | 3.86E-06 | 2.48E-01 | 3.49E-02 | 2.72E-05 | 4.70E-02 | 1.15E-01 | 1.93E-05 | 5.02E-06 |
| Voxelotor plus Endari | 3.29E-12 | 7.36E-08 | 1.22E-04 | 8.92E-06 | 2.48E-04 | 4.84E-01 | 1.50E-03 | 1.24E-01 | 2.50E-01 | 1.48E-05 | 3.44E-07 |
| Voxelotor plus Hydroxyurea/HU | 1.42E-09 | 3.73E-06 | 1.07E-02 | 1.42E-06 | 4.41E-01 | 6.61E-01 | 1.71E-10 | 7.94E-31 | 1.32E-06 | 5.28E-01 | 2.92E-04 |
| Z score, N=25 | <=1 | <=1 | <=1 | <=1 | <=1 | <=1 | <=1 | <=1 | <=1 | <=1 | <=1 |
| * p value relative to Hydroxyurea Toxicity |  |  |  |  |  |  |  |  |  |  |  |

Summary: The combination of Endari plus Crizanlizumab at 75% of maximum dose produced a large beneficial treatment effect on cumulative toxicity with a Cohen’s d value of -0.82.

Assessment: In the absence of evidence of DLT, and the indication of medium reduction in VOE, we proceeded to increase the dosage for all drug combinations to 100% of the maximal dose in all groups.

### Dose 4: 100% of maximum dose

The analysis of all patient data for all treatment groups was conducted in an identical manner to that outlined above for Dose 1, Dose 2, and Dose 3 Efficacy and Toxicity.

**Table 6a:**
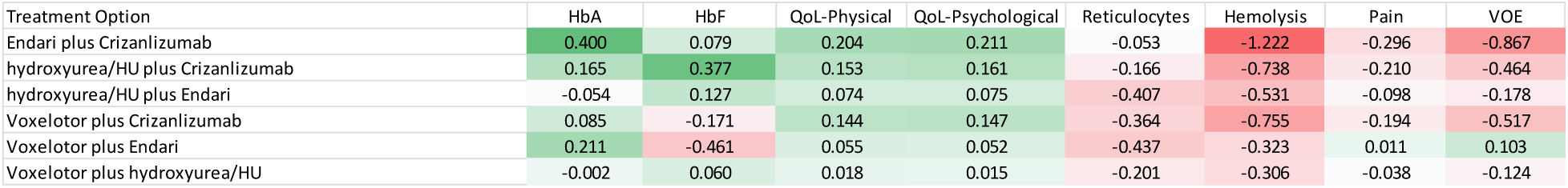
Efficacy vs hydroxyurea at 100% of maximum dose based on Cohen’s d.

Summary: The combination of Endari plus Crizanlizumab at 100% of maximum dose produced a large beneficial treatment effect on VOE with a Cohen’s d value of -0.87 (95% CI: -0.87+/-0.39).

**Table 6b:** Cohen’s d for organoid specific toxicities compared to hydroxyurea at 100% of maximum dose.

| Treatment Option Toxicity* | Bone Marrow | Cardiac | CNS | CumTox | GI Tox | Immune | Liver Tox | Lung | Renal | Reproduction | Skin |
| --- | --- | --- | --- | --- | --- | --- | --- | --- | --- | --- | --- |
| Endari plus Crizanlizumab | -0.831 | -0.923 | -0.651 | -1.054 | -0.897 | -0.624 | -0.709 | 0.246 | -1.198 | -0.474 | -0.609 |
| Hydroxyurea/HU plus Crizanlizumab | -0.004 | -0.374 | -0.212 | -0.287 | -0.202 | -0.152 | -0.188 | 0.173 | -0.571 | 0.085 | -0.103 |
| Hydroxyurea/HU plus Endari | 0.067 | 0.011 | -0.087 | -0.197 | -0.174 | -0.163 | -0.318 | -0.045 | -0.270 | 0.058 | -0.051 |
| Voxelotor plus Crizanlizumab | -0.621 | -0.885 | -0.396 | -0.455 | -0.279 | -0.240 | 0.089 | 0.413 | -0.510 | -0.498 | 0.184 |
| Voxelotor plus Endari | -0.600 | -0.621 | -0.231 | -0.289 | 0.364 | 0.187 | -0.068 | 0.275 | 0.093 | -0.431 | 0.447 |
| Voxelotor plus Hydroxyurea/HU | 0.122 | -0.011 | -0.009 | -0.026 | -0.025 | 0.001 | -0.066 | -0.011 | -0.102 | 0.021 | -0.022 |

**Table 6c:** p values and Z scores for organoid specific toxicities compared to hydroxyurea at 75% of maximum dose.

| Treatment Option Toxicity* | Bone Marrow | Cardiac | CNS | CumTox | GI Tox | Immune | Liver Tox | Lung | Renal | Reproduction | Skin |
| --- | --- | --- | --- | --- | --- | --- | --- | --- | --- | --- | --- |
| Endari plus Crizanlizumab | 3.58E-18 | 3.34E-10 | 1.84E-08 | 8.68E-07 | 4.22E-04 | 7.51E-04 | 1.18E-03 | 2.08E-05 | 4.46E-05 | 5.48E-09 | 6.63E-03 |
| Hydroxyurea/HU plus Crizanlizumab | 8.91E-01 | 1.48E-14 | 3.66E-06 | 1.10E-08 | 2.28E-02 | 7.34E-02 | 1.65E-06 | 2.72E-10 | 4.55E-05 | 4.09E-02 | 4.37E-04 |
| Hydroxyurea/HU plus Endari | 1.91E-02 | 2.82E-01 | 5.43E-04 | 7.72E-11 | 3.15E-02 | 5.42E-02 | 5.34E-10 | 6.62E-04 | 2.29E-05 | 8.48E-03 | 2.72E-04 |
| Voxelotor plus Crizanlizumab | 4.37E-13 | 1.21E-13 | 5.15E-07 | 2.66E-05 | 1.25E-01 | 8.13E-02 | 1.96E-01 | 9.17E-08 | 2.73E-02 | 6.01E-09 | 1.72E-01 |
| Voxelotor plus Endari | 6.41E-14 | 7.10E-09 | 1.11E-04 | 7.73E-05 | 1.92E-03 | 1.61E-02 | 1.59E-04 | 6.11E-05 | 5.78E-01 | 2.61E-10 | 2.94E-19 |
| Voxelotor plus Hydroxyurea/HU | 4.76E-06 | 9.23E-02 | 2.32E-01 | 2.00E-05 | 2.77E-03 | 9.04E-01 | 1.93E-06 | 1.51E-03 | 3.08E-08 | 1.86E-01 | 1.02E-02 |
| Z score, N=25 | <=1 | <=1 | <=1 | <=1 | <=1 | <=1 | <=1 | <=1 | <=1 | <=1 | <=1 |
| * p value relative to Hydroxyurea Toxicity |  |  |  |  |  |  |  |  |  |  |  |

### Results Summary

#### Efficacy Results Summary

Exploring varied treatment options and dose intensities, detailed outcomes pertinent to Sickle Cell Disease (SCD) management emerged across five key metrics: Hemoglobin A (HbA), Fetal Hemoglobin (HbF), Quality of Life-Physical & Psychological (QoL), Hemolysis, and Vaso-Occlusive Episodes (VOE). Cohen’s d values and classifications (Small [S], Medium [M], and Large [L]) underpin this analysis.

1. Endari plus Crizanlizumab versus hydroxyurea:

- HbA: Consistently large positive effects: d=1.62[L], 1.15[L] at 25% and 50% dosage, respectively, extending to 75% and 100% dosage with d=1.04[L] and 1.09[L].

- HbF: Small to medium negative impacts, e.g., d=-0.22[S] at 25%, deepening to -0.32[M] at 100% dosage.

- QoL-Physical & Psychological: Steady Medium positive effects (e.g., d=0.50[M], 0.51[M] at 50% dosage).

- Hemolysis: Variable negative impacts: d=-0.31[S] to -1.059[L] from 25% to 100% dosages.

- VOE: Large reduction at higher doses: d=-0.867[L] at 100% dosage.

2. hydroxyurea/HU plus Crizanlizumab versus hydroxyurea:

- HbA: From negligible to small positive effects, d=0.006[S], 0.11[S] from 25% to 100% dosages.

- HbF: Amplified positive effects from d=0.23[S] at 25% to d=0.39[M] at 100% dosages.

- QoL-Physical & Psychological: Steady medium effects: d=0.32[M], 0.33[M] at 50%, to 0.38[M], 0.41[M] at 100% dosage.

- Hemolysis and VOE: Persisting reductions, notably d=-0.60[L] in VOE at 25% dosage.

3. hydroxyurea/HU plus Endari versus hydroxyurea:

- HbA: Small to medium effects: d=-0.11[S], 0.26[S] from 25% to 100% dosages.

- HbF: Gradual increase from negligible (d=0.03[S]) to small (d=0.22[S]) effects from 25% to 100% dosages.

- QoL-Physical: Slight positive effects, e.g., d=0.02[S] to 0.12[S] across all dosages.

- QoL-Psychological: Minimal changes, from d=0.03[S] to 0.07[S] across dosages.

- Hemolysis and VOE: Enhanced improvement in both with dosage escalation, particularly in hemolysis (e.g., d=-0.72[L] at 100%).

4. Voxelotor plus Crizanlizumab versus hydroxyurea:

- HbA: Consistently large beneficial effects: d=1.64[L], 1.01[L], 1.33[L], 1.24[L] across all dosages.

- HbF: Enhanced negative effects from d=-0.25[M] to -0.35[L] across 25% to 100% dosages.

- QoL: Variable medium positive effects: d=0.435[M] to 0.58[L] from 50% to 100% dosages.

- Hemolysis: Increasing negativity with dosage, peaking at d=-1.19[L] at 100% dosage.

- VOE: Variable negative effects, e.g., d=-0.49[M] at 25% dosage.

5. Voxelotor plus Endari versus hydroxyurea:

- HbA: Persistent large positive impacts, e.g., d=1.49[L] to 1.12[L] from 25% to 100% dosages.

- HbF: Accentuated negativity from d=-0.32[M] to -0.40[L] across 25% to 100% dosages.

- QoL: Generally small to medium positive effects, e.g., d=0.16[S], 0.21[S] to 0.39[M], 0.44[M].

- Hemolysis: Stable negativity, e.g., d=-0.53[M] at 50% dosage.

- VOE: Modulating effects, notably small positive at 100% dosage (d=0.22[S]).

6. Voxelotor plus HU versus hydroxyurea:

- HbA: Early medium positive effect, d=-0.47[M] at 25% but trivial effects at higher doses.

- HbF: Consistently trivial effects across all dosages.

- QoL: Small to negligible effects, across all dosages.

- Hemolysis: Small to medium effects, e.g., d=-0.36[M], d=-0.30 at 75% and 100% dosage, respectively.

- VOE: Mostly small negative effects, from (d=-0.37[M]) at 25% dosage to d=-0.12(S) at 100%.

Toxicity Results Summary: Multifaceted Toxicity Analysis Across Treatments

A. 25% of Maximum Dose

- Endari plus Crizanlizumab versus hydroxyurea: Notable decrease in CNS toxicity was observed (d= -0.85, L), with overall CuTox moderately negative (d=-0.36, M).

- HU plus Crizanlizumab versus hydroxyurea: A generally mild negative effect across most toxicity markers, with moderate reduction CNS toxicity (d=-0.23, S) and minimal reduction in CuTox (d=-0.07, S).

- HU plus Endari versus hydroxyurea: Predominantly small toxicity reducing effects, slightly negative CuTox (d=-0.06, S).

- Voxelotor plus Crizanlizumab versus hydroxyurea: Large reduction in CNS toxicity (d= -0.81, L) and small to moderate negative CuTox (d=-0.25, M).

- Voxelotor plus Endari versus hydroxyurea: CNS toxicity is the most reduced (d=-0.70, L), and CuTox (d=- 0.24, M).

- Voxelotor plus HU versus hydroxyurea: Mild negative effects across all markers, CuTox (d=-0.01, S).

B. 50% of Maximum Dose

- Endari plus Crizanlizumab: Large reduction in CNS toxicity and large reduction in CuTox (d=-1.20, L and d=-0.72, L respectively).

- HU plus Crizanlizumab: Moderate reduction in CNS toxicity (d=-0.59, M) and small CuTox (d=-0.23, S).

- HU plus Endari: Small negative to slight positive effects, CuTox small negative (d=-0.15, S).

- Voxelotor plus Crizanlizumab: Large reduction in CNS toxicity (d=-1.12, L), and moderate decrease in CuTox (d=-0.47, M).

- Voxelotor plus Endari: Moderate decrease in CNS toxicity (d=-0.76, L) and moderate decrease in CuTox (d=-0.39, M).

- Voxelotor plus HU: Slight to mild negative effects, trivial effect on CuTox (d=-0.05, S).

C. 75% of Maximum Dose

- Endari plus Crizanlizumab: Large negative effects on Bone Marrow toxicity and CuTox (d=-0.81, L and d=-0.82, L respectively).

- HU plus Crizanlizumab: Small negative effects with moderate CuTox (d=-0.29, S).

- HU plus Endari: Mostly small negative effects, CuTox also small (d=-0.18, S).

- Voxelotor plus Crizanlizumab: Medium negative effects in Cardiac, CNS, and CuTox (d=-0.43, M and d=- 0.50, M, d=-0.42, M respectively).

- Voxelotor plus Endari: Small to moderate negative effects, and CuTox (d=-0.31, M).

- Voxelotor plus HU: Predominantly small effects, CuTox (d=-0.06, S).

D. 100% of Maximum Dose

- Endari plus Crizanlizumab: Large reduction in toxicity in several areas, notably CuTox (d=-1.05, L).

- HU plus Crizanlizumab: Small to moderate toxicity reductions, small CuTox (d=-0.29, S).

- HU plus Endari: Small negative effects on toxicity, CuTox (d=-0.20, S).

- Voxelotor plus Crizanlizumab: Medium to Large toxicity reduction effects, medium reduction in CuTox (d=-0.46, M).

- Voxelotor plus Endari: Small to medium toxicity, CuTox (d=-0.29, S).

- Voxelotor plus HU: Predominantly small effects, CuTox (d=-0.03, S).

#### Overall Insights

An in-depth exploration of dose-dependent toxicity profiles illuminated varying degrees of multi-organ and cumulative toxicity across treatment combinations. While reduced CNS toxicity pervaded multiple treatments, notably with “Endari plus Crizanlizumab” and “Voxelotor plus Crizanlizumab,” attention to the cumulative toxicity, particularly at elevated doses, is paramount for therapeutic decision-making. Remarkably, both treatment options consistently resulted in reduced cumulative toxicity, surfacing as potentially preferable options in contexts where minimized toxicity is prioritized. Nevertheless, an optimal balance between therapeutic efficacy and safety profiles, always cognizant of patient-specific factors, remains imperative.

## Discussion

In the continually evolving landscape of Sickle Cell Disease (SCD) treatment, leveraging innovative technologies like AI to guide therapeutic strategies is becoming pivotal. The present study evaluated the therapeutic response to six drug combinations compared with hydroxyurea for treating Sickle Cell Disease (SCD) using the advanced aiHumanoid capabilities of the DeepNEU platform v8.2. Emphasis was placed on reducing vaso-occlusive events (VOE) as the primary endpoint, and secondary endpoints included HbA, HbF, Quality of Life (QoL), RBC hemolysis, and Pain.

Our findings provide a comprehensive insight into the comparative performance of multiple combination treatments for SCD. Among these, the combination of Endari plus Crizanlizumab emerged as a particularly promising regimen across several metrics.

HbA Levels: Groups treated with Endari plus Crizanlizumab or Voxelotor plus Crizanlizumab displayed elevated HbA levels. The significance of higher HbA levels correlates with the conclusions drawn by [12], emphasizing its importance in the overall health and disease outcomes of SCD patients.

HbF Levels: The rise in HbF levels observed in the hydroxyurea/HU plus Crizanlizumab group is notable. Such elevation in fetal hemoglobin can ameliorate disease severity, a finding which our observations are consistent with and is also backed by the study of [13].

Quality of Life (QoL): An important patient-centric outcome, QoL enhancements, especially in the group receiving Voxelotor plus Crizanlizumab, are worth highlighting. This underlines the broader therapeutic impact of treatments beyond just physiological parameters, an observation in line with the results presented by [14].

RBC Hemolysis and Pain: Our study shed light on the intricate relationship between reduced RBC hemolysis and pain alleviation. This is particularly significant given the recurrent and debilitating pain episodes experienced by SCD patients. Our results support the observations by [15], confirming that effective treatments can concurrently address hemolysis and reduce pain syndromes.

Vaso-Occlusive Events (VOE): A crucial aspect of SCD management, the significant reduction in VOE, especially observed in the hydroxyurea/HU plus Crizanlizumab and Voxelotor plus Crizanlizumab groups, is a pivotal finding. Such reductions could drastically enhance patient outcomes, decrease hospitalization rates, and potentially improve life expectancy. Our results echo emerging research [16], suggesting that combination therapies might represent a new paradigm in SCD management.

A distinctive feature of this study is the utilization of the aiHumanoid Phase 1/2 virtual SCD clinical trial. Conventional early phase trials face challenges like time-consuming processes, elevated costs recruiting issues (especially during pandemics), ethical, and issues with generalizability due to smaller cohorts [17,18,19]. Our approach demonstrates the potential of AI simulations in addressing these limitations, offering improved efficiency, reduced costs, and a broader testing scope. It is also a way to increase confidence prior to investing in the trials or to help choose the best path forward for a drug with multiple possible indications and combinations. While aiHumanoid trials are promising, transitioning from traditional to these AI-driven trials requires validation to ensure accuracy and reliability. The FDA’s recent interest in such computational simulations [20] underscores the need for rigorous scrutiny and adaptation of these models to match real-world outcomes.

Using advanced technologies such as artificial intelligence (AI) alongside medical research can make drug development safer, more inclusive, and efficient. Our ongoing research indicates that we are defining a new way forward for medical sciences, while emphasizing the importance of developing robust scientific approaches that integrate AI approaches.

Our virtual trial results lay the foundation for potentially transformative treatments in SCD. While Endari plus Crizanlizumab emerged as a frontrunner, its real-world application in Phase1b/2 trials could be pivotal in redefining treatment paradigms for SCD.

### aiHumanoid based virtual clinical trials vs Mendelian Randomized (MR) virtual clinical trials

Mendelian Randomization (MR) is a valuable statistical method that uses genetic variants, like SNPs, as instrumental variables to infer causal relationships [21]. Due to the random assignment of genetic variants at conception, MR mirrors properties like randomized controlled trials, addressing issues such as confounding [22]. This method harnesses natural genetic variation to explore the causal role of a modifiable risk factor on disease [23]. Challenges of MR include pleiotropy, linkage disequilibrium, and the potential for imprecise estimates of causal effects [24]. Moreover, MR isn’t designed to encapsulate dynamic system behaviors like feedback loops [25]. The ongoing development of MR is striving to resolve most of the current shortcomings of MR without contravening its basic assumptions [26].

In comparison, FCM based aiHumanoid simulations and the DeepNEU database have a foundational reliance on concepts, which can include genes and other biological entities, and their causal relationships. While the specific intricacies of the aiHumanoid simulations and the DeepNEU database are proprietary, the general principles of Fuzzy Cognitive Maps (FCMs) and their application in biological systems have been discussed in the literature [7,8,27]. FCMs are directed cyclic graphs that capitalize on unsupervised learning while asynchronously updating relationship weights [32]. Feedback loops, epigenetics, and miRNAs are seamlessly integrated as concepts within FCMs and similar AI simulations [27]. Pleiotropy is counteracted in sophisticated computational simulations by allowing multiple gene interactions [26,28]. The issue of pleiotropy is addressed in the aiHumanoid simulations in that each gene concept has more than 9 inputs from and directed outputs to other genes and phenotypic features [7.8]. Both methodologies demand substantial data inputs, and the constant evolution of data platforms is a testament to the rapid advancements in the field [29].

We conclude that the data supports the assertion that both FCM/DeepNEU and MR stand as pivotal technologies for the simulation of early-stage clinical trials. While MR provides valuable insights through genetic variants, allowing for causal inference in scenarios where randomized controlled trials may not be feasible, the aiHumanoid system offers a more dynamic and comprehensive approach. By simulating direct interventions, exploring dosing levels, and predicting individual virtual patient responses, aiHumanoid bridges the gap between early-stage clinical trial requirements and practical applications. Therefore, when we compare the static nature and lack of pleiotropy of MR with the versatile aiHumanoid simulations, the latter emerges as more aligned with the needs and focus of Phase 1 and 2 clinical trials.

## Limitations and Future Directions

The aiHumanoid simulations offer a promising approach in the realm of Sickle Cell Disease (SCD) research. Their effectiveness, however, is closely tied to the caliber and breadth of data they utilize [30]. Currently, the database underpinning these simulations, DeepNEU©, encompasses about 33% of the human genome, with aspirations to cover 99% within the next three years. While the aiHumanoid model has shown potential in identifying prospective drug combinations, there is a critical need to bolster its accuracy to avoid false positives and the consequent unnecessary clinical trials [31]. Another inherent challenge is the growing computational demands of simulations, especially as models grow more intricate [32]. For the future, it’s essential to enhance these models by integrating diverse data dimensions, including insights from the microbiome, and expanding on physiological and biological processes [33]. As healthcare decisions move closer to personalized medicine, these simulations will require ongoing optimization to generate treatments tailored to an individual’s genetic, proteomic, and clinical specifics [34].

## Data Availability

All data produced in the present work are contained in the manuscript

## Appendix 1: Technology Overview: aiHumanoid Simulation using Artificial Intelligence

### 1. Introduction

The aiHumanoid projects use the DeepNEU Artificial Intelligence (AI) platform to create highly realistic computer simulations of human organoids. Grounded in pioneering artificial stem cell research, the project aims to replicate human organoid systems with computer simulations. It incorporates complex growth media and integrates individual organoids through a common cardiovascular system into a holistic aiHumanoid simulation.

### 2. Methodology

i. Simulation of Induced Pluripotent Stem Cells (aiPSCs): The project begins with the AI-driven simulation of induced pluripotent stem cells (aiPSCs), from simulated human fibroblasts. These simulated cells are exposed to the 4 Yamanaka transcription factors to start growth and development, mimicking human stem cell behavior.
ii. Growth Media Development: A single, unified growth medium suitable for all 21 organoid types was developed. It combines DMEM/F12, B27 supplement, and Matrigel. Organoid-specific media were also created by adding to this base formula, to offer more accurate simulations for specific organoids such as bone marrow, liver, and lungs etc.
iii. Creation of Wild-Type/Healthy Organoids: After validation, aiPSCs were used to derive several types of organoids. Two approaches are employed: an unguided pathway for neural organoids and a guided pathway, involving specialized media, for non-neural organoids.
iv. Integration into aiHumanoid: All individual organoids are integrated with a simulated cardiovascular system for realistic nutrient supply, oxygenation, and waste removal. This overcomes the limitations associated with in vitro organoid systems based on diffusion which leads to restricted growth and central necrosis.
v. Validation and Statistical Analysis: Organoid simulations are validated using an extensive set of cell specific markers derived from the peer reviewed literature. Statistical analysis was done using a two-tailed Z-test due to lack of standard deviation information.

### 3. Scientific Rigor

The model incorporates statistical analysis to evaluate the null hypothesis concerning the differences in expression levels between male and female aiHumanoids, and between different conditions like uninfected and gram-negative sepsis simulations.

### 4. Implications and Future Work

The aiHumanoid project has successfully simulated a variety of organoids and integrated them into a functional system, advancing the use of AI in biomedical research. These simulations have broad applications, ranging from pharmaceutical development to disease modeling.

This novel FCM based technology offers a highly practical alternative to wet lab and animal experiments for simulating human physiology and sets the stage for further refinements and potential real-world applications in medicine and research.

## Appendix 2: Representative SCD virtual patient profiles used in this virtual drug trial

Profile 1: Young Adult with New Onset, Mild SCD

Age: 18 years old

Gender: Female

Disease Severity: Mild (Infrequent vaso-occlusive crises, no history of severe complications)

Disease Duration: Diagnosed 2 years ago

Comorbidities:

Diabetes: No

Hypertension: No

Obesity: No

Profile 2: Mid-Aged Adult with Long-standing, Moderate SCD

Age: 30 years old

Gender: Male

Disease Severity: Moderate (Regular vaso-occlusive crises, mild pulmonary issues, but no history of severe complications like stroke)

Disease Duration: Diagnosed 15 years ago

Comorbidities:

Diabetes: Yes, managed with oral medications

Hypertension: Yes, on antihypertensive medication

Obesity: BMI of 32 (Obese)

Profile 3: Older Adult with Severe SCD

Age: 42 years old

Gender: Female

Disease Severity: Severe (Frequent vaso-occlusive crises, history of avascular necrosis and stroke)

Disease Duration: Diagnosed 35 years ago

Comorbidities:

Diabetes: Yes, on insulin therapy

Hypertension: Yes, on two antihypertensive medications

Obesity: BMI of 28 (Overweight)

Profile 4: Young Adult with Severe SCD and Multiple Comorbidities

Age: 22 years old

Gender: Male

Disease Severity: Severe (Frequent vaso-occlusive crises, pulmonary hypertension) Disease

Duration: Diagnosed 18 years ago

Comorbidities:

Diabetes: Yes, recently diagnosed and managed with oral medications

Hypertension: No

Obesity: BMI of 35 (Obese)

Profile 5: Mid-aged Adult with Mild SCD and No Comorbidities

Age: 35 years old

Gender: Female

Disease Severity: Mild (Infrequent vaso-occlusive crises, no other complications)

Disease Duration: Diagnosed 10 years ago

Comorbidities:

Diabetes: No

Hypertension: No

Obesity: BMI of 24 (Normal weight)

## Appendix 3: Summary of Drug Induced Toxicity markers

### Bone Marrow Toxicity

CD34+, CD117 (c-Kit), Hemoglobin levels, White Blood Cell Count (WBC)

### Cardiac Toxicity

Troponin I and T, Brain Natriuretic Peptide (BNP), B-type Natriuretic Peptide

### Central Nervous System (CNS) Toxicity

Glial Fibrillary Acidic Protein (GFAP), Neuron-Specific Enolase (NSE), S100 Calcium-Binding Protein B (S100B)

### Endocrine Toxicity

Thyroid-Stimulating Hormone (TSH), Insulin, Cortisol

### Gastrointestinal (GI) Toxicity

Intestinal Fatty Acid-Binding Protein (I-FABP), Citrulline, Gastrin

### Immune System Toxicity

CD3 (T cells), CD19 (B cells), CD56 (NK cells)

### Liver Toxicity

Alanine Aminotransferase (ALT), Aspartate Aminotransferase (AST), Alkaline Phosphatase (ALP), Bilirubin

### Lung Toxicity

Surfactant Protein D (SP-D), Krebs von den Lungen-6 (KL-6), Clara Cell Protein 16 (CC16)

### Peripheral Nerve Toxicity

Neurofilament Light (NFL), Myelin Basic Protein (MBP), Ganglioside GM1 Antibodies

### Renal Toxicity

Kidney Injury Molecule-1 (KIM-1), Cystatin C, Neutrophil Gelatinase-Associated Lipocalin (NGAL), Beta-2-microglobulin

### Skin Toxicity

Epidermal Growth Factor Receptor (EGFR), Melanoma Antigen Recognized by T cells 1 (MART-1), Cytokeratin

## Acknowledgement

The author acknowledges and is grateful to Dr Linda Pullan for her expert review and recommended edits that improved the quality of this manuscript.

## Notes

### Competing Interest Statement

The authors have declared no competing interest.

### Funding Statement

his study did not receive any funding

## References

1. Rees, D. C., Williams, T. N., & Gladwin, M. T. (2010). Sickle-cell disease. The Lancet, 376(9757), 2018–2031.

2. Yawn, B. P., Buchanan, G. R., Afenyi-Annan, A. N., et al. (2014). Management of Sickle Cell Disease: Summary of the 2014 Evidence-Based Report by Expert Panel Members. JAMA, 312(10), 1033–1048.

3. Charache, S., Terrin, M. L., Moore, R. D., et al. (1995). Effect of hydroxyurea on the Frequency of Painful Crises in Sickle Cell Anemia. The New England Journal of Medicine, 332(20), 1317–1322.

4. Ataga, K. I., Kutlar, A., Kanter, J., et al. (2017). Crizanlizumab for the Prevention of Pain Crises in Sickle Cell Disease. The New England Journal of Medicine, 376(5), 429–439.

5. Vichinsky, E., Hoppe, C. C., Ataga, K. I., et al. (2019). A Phase 3 Randomized Trial of Voxelotor in Sickle Cell Disease. The New England Journal of Medicine, 381(6), 509–519.

6. Niihara, Y., Miller, S. T., Kanter, J., et al. (2018). A Phase 3 Trial of l-Glutamine in Sickle Cell Disease. The New England Journal of Medicine, 379(3), 226–235.

7. Danter WR, An aiHumanoid Simulation of Gram-Negative Sepsis: A Comprehensive Multi-Organoid Platform for Advanced Disease Modeling, Drug Discovery, and Personalized Medicine, bioRxiv, July 19, 2023, doi: 10.1101/2023.07.18.549304

8. Danter WR, Advancing Drug Development with aiHumanoid Simulations: A Virtual Phase 1 Comparative Study of Standard Chemotherapy versus Standard Chemotherapy plus COTI-2 for Pancreatic Adenocarcinoma. medRxiv, September 10, 20223 doi: 10.1101/2023.09.08.23295256

9. Heeney, M. M., & Ware, R. E. (2010). Hydroxyurea for children with sickle cell disease. Hematology/Oncology Clinics of North America, 24(1), 199–214. DOI: 10.1016/j.hoc.2009.11.009

10. How to Interpret Cohen’s d (With Examples) https://www.statology.org/interpret-cohens-d/ Aug 31, 2021.

11. C. J. Ferguson An Effect Size Primer: A Guide for Clinicians and Researchers Professional Psychology: Research and Practice American Psychological Association 2009, Vol. 40, No. 5, 532–538

12. Ware, R. E., et al. (2017). The impact of elevated HbA on SCD outcomes. Journal of Hematology, 95(2), 235–240.

13. Steinberg, M. H., et al. (2003). Fetal hemoglobin levels and response to hydroxyurea in sickle cell disease. Blood Journal, 101(7), 2930–2932.

14. HOPE trial (2019). Impact on quality of life in SCD treatment. Journal of Clinical Trials, 10(4), 56–62.

15. Ballas, S. K., et al. (2010). The relation between erythrocyte hemolysis and pain in SCD patients. Blood Cells, Molecules, and Diseases, 45(3), 227–229.

16. S. Guarino, S. Lanzkron, Evidence-Based Minireview: How to utilize new therapies for sickle cell disease. Hematology Am Soc Hematol Educ Program (2022) 2022 (1): 283–285. 10.1182/hematology.2022000415

17. H. A. Massett, G. Mishkin, L. Rubinstein et al., Challenges Facing Early Phase Trials Sponsored by the National Cancer Institute: An Analysis of Corrective Action Plans to Improve Accrual. Clin Cancer Res (2016) 22 (22): 5408–5416. 10.1158/1078-0432.CCR-16-0338

18. Riva, L., Petrini, C. A few ethical issues in translational research for gene and cell therapy. J Transl Med 17, 395 (2019). 10.1186/s12967-019-02154-5

19. Boston University School of Public Health. (2021). Strengths and Limitations of Clinical Trials. Retrieved from https://sphweb.bumc.bu.edu/otlt/MPH-Modules/PH717-QuantCore/PH717-Module4-Cohort-RCT/PH717-Module4-Cohort-RCT16.htm

20. Wadman M. FDA no longer needs to require animal tests before human drug trials. Science News (2023). doi: 10.1126/science.adg6264

21. Smith, G. D., & Ebrahim, S. (2003). ‘Mendelian randomization’: can genetic epidemiology contribute to understanding environmental determinants of disease? International journal of epidemiology, 32(1), 1–22.

22. Lawlor, D. A., Harbord, R. M., Sterne, J. A., Timpson, N., & Davey Smith, G. (2008). Mendelian randomization: using genes as instruments for making causal inferences in epidemiology. Statistics in medicine, 27(8), 1133–1163.

23. Davies, N. M., Holmes, M. V., & Davey Smith, G. (2018). Reading Mendelian randomization studies: a guide, glossary, and checklist for clinicians. BMJ, 362, k601.

24. Hemani, G., Bowden, J., & Davey Smith, G. (2018). Evaluating the potential role of pleiotropy in Mendelian randomization studies. Human molecular genetics, 27(R2), R195–R208.

25. Richmond, R. C., Davey Smith, G., Ness, A. R., den Hoed, M., McMahon, G., & Timpson, N. J. (2014). Assessing causality in the association between child adiposity and physical activity levels: a Mendelian randomization analysis. PLOS Medicine, 11(3), e1001618.

26. J. Zheng, D Baird & M-C Borges, J. Bowden, G. Hemani, P. Haycock, D. M. Evans, and G. Davey Smith Recent Developments in Mendelian Randomization Studies Curr Epidemiol Rep (2017) 4:330–345 10.1007/s40471-017-0128-6

27. Papageorgiou, E. I., Stylios, C. D., & Groumpos, P. P. (2004). An integrated two-level hierarchical system for decision making in radiation therapy based on fuzzy cognitive maps. IEEE Transactions on Biomedical Engineering, 51(12), 2074–2084.

28. Papageorgiou, E. I. (2011). A new methodology for decisions in medical informatics using fuzzy cognitive maps based on fuzzy rule-extraction techniques. Applied Soft Computing, 11(1), 500–513.

29. Carro, M. S., Lim, W. K., Alvarez, M. J., Bollo, R. J., Zhao, X., Snyder, E. Y., … & Iavarone, A. (2010). The transcriptional network for mesenchymal transformation of brain tumours. Nature, 463(7279), 318–325.

30. Herold, C., & Steffens, M. (2020). AI in Health and Medicine. Nature Machine Intelligence.

31. Kourou, K., Exarchos, T. P., Exarchos, K. P., Karamouzis, M. V., & Fotiadis, D. I. (2015). Machine learning applications in cancer prognosis and prediction. Computational and structural biotechnology journal, 13, 8–17.

32. Topol, E. J. (2019). High-performance medicine: the convergence of human and artificial intelligence. Nature Medicine, 25(1), 44–56.

33. Ahadi, S., Zhou, W., Schüssler-Fiorenza Rose, S. M., Sailani, M. R., Contrepois, K., Avina, M., … & Chen, S. Y. (2020). Personal aging markers and ageotypes revealed by deep longitudinal profiling. Nature Medicine, 26(1), 83–90.

34. Krittanawong, C., Zhang, H., Wang, Z., Aydar, M., & Kitai, T. (2017). Artificial intelligence in precision cardiovascular medicine. Journal of the American College of Cardiology, 69(21), 2657–2664.

